# Decreased Immune Response to COVID-19 mRNA Vaccine in Patients with Inflammatory Bowel Diseases Treated with Anti TNFα

**DOI:** 10.1101/2021.08.22.21262263

**Authors:** Hadar Edelman-Klapper, Eran Zittan, Ariella Bar-Gil Shitrit, Keren Masha Rabinowitz, Idan Goren, Irit Avni-Biron, Jacob E. Ollech, Lev Lichtenstein, Hagar Banai-Eran, Henit Yanai, Yifat Snir, Maor H. Pauker, Adi Friedenberg, Adva Levy-Barda, Arie Segal, Yelena Broitman, Eran Maoz, Baruch Ovadia, Maya Aharoni Golan, Eyal Shachar, Shomron Ben-Horin, Tsachi-Tsadok Perets, Rami Eliakim, Sophy Goren, Michal Navon, Noy Krugliak, Michal Werbner, Joel Alter, Moshe Dessau, Meital Gal-Tanamy, Natalia T. Freund, Dani Cohen, Iris Dotan, On behalf of the REsponses to COVid-19 vaccinE IsRaeli IBD group [RECOVERI]

**Author notes:** Corresponding author: Prof. Iris Dotan, MD. Division of Gastroenterology, Rabin Medical Center, Beilinson Hospital, Jabotinsky St. 39, Petah Tikva, 49100, Israel.

## Abstract

**Background:** Patients with inflammatory bowel diseases (IBD), specifically those treated with anti-tumor necrosis factor (TNF)α biologics are at high risk for vaccine preventable infections. Their ability to mount adequate vaccine responses is unclear.

**Aim:** to assess immune responses to mRNA-COVID-19 vaccine, and safety profile, in patients with IBD stratified according to therapy, compared to healthy controls (HC).

**Methods:** Prospective, controlled, multi-center Israeli study. Subjects enrolled received two BNT162b2 (Pfizer/BioNTech) doses. Anti-spike (S) antibodies levels and functional activity, anti-TNFα levels and adverse events (AEs) were detected longitudinaly.

**Results:** Overall 258 subjects: 185 IBD (67 treated with anti-TNFα), and 73 HC. After the first vaccine dose all HC were seropositive, while some patients with IBD, regardless of treatment, remained seronegative. After the second dose all subjects were seropositive, however anti-S levels were significantly lower in anti-TNFα treated compared to untreated patients, and HC (p<0.001; p<0.001, respectively). Neutralizing and inhibitory functions were both lower in anti-TNFα treated compared to untreated patients, and HC (p<0.03; p<0.0001, respectively). Anti-TNFα drug levels and vaccine responses did not affect anti-S levels. Infection rate (∼2%) and AEs were comparable in all groups. IBD activity did not change in response to BNT162b2.

**Conclusions:** In this prospective study in patients with IBD stratified according to treatment all patients mounted an immune response to two doses of BNT162b2. However, its magnitude was significantly lower in patients treated with anti-TNFα, regardless of administration timing and drug levels. Vaccine was safe. As vaccine immune response longevity in this group may be limited, vaccine booster dose should be considered.

## Introduction

Severe acute respiratory syndrome coronavirus 2 (SARS-CoV-2) infection and coronavirus disease 2019 (COVID-19) resulted in a worldwide pandemic^1^. To face the immense morbidity and mortality burden, accelerated vaccine development programs and mass vaccination campaigns were conducted. Vaccine studies included healthy adults or those with stable chronic diseases^2, 3^. Patients with inflammatory bowel diseases (IBD), both Crohn’s disease (CD) and ulcerative colitis (UC) were excluded. These patients are often treated with immunomodulators and/or biologic therapy such as anti-tumor necrosis factor (TNF) α, potentially associated with an increased risk of infection^4–6^. While guidelines recommend vaccination per standard immunization schedules^4, 7, 8^, patients’ ability to mount an adequate immune response to certain vaccines or infections is doubted^6, 9–17^. This was even less clear for the new mRNA-based vaccines against SARS-CoV-2. Concerns regarding adverse events (AEs), including IBD exacerbation, further underscored the need for vaccine responses assessment in these patients.

A massive vaccination campaign against COVID-19 started in Israel on December 19, 2020, with mRNA-based COVID-19 vaccine (BNT162b2, Pfizer/BioNTech), administered in two doses three weeks apart^18^. We conducted a prospective multi-center Israeli study to assess immune responses to BNT162b2 in patients with IBD stratified according to therapy, compared to healthy controls (HC).

## Methods

### Study design and participants

A prospective, observational, multi-center study was conducted to assess immune responses to the mRNA-based COVID-19 vaccine BNT162b2, their dynamics, predictors of response and safety, in patients with IBD compared to HC. A call for patient referral was distributed to all Israeli gastroenterologists and patients with IBD on December 28, 2020. Patients aged≥18 years were recruited. IBD diagnosis was defined by accepted criteria. HC group included volunteers (healthcare professionals and their relatives) without known gastrointestinal diseases. Patients with past COVID-19 infection proved by SARS-CoV-2 polymerase-chain-reaction test and pregnant women were excluded. Patients with IBD were stratified at baseline into those treated with anti-TNFα, or those with any other IBD treatment or untreated. All participants received two 30µg BNT162b2 vaccine doses intramuscularly, administered 21-28 days apart, as per manufacturers’ recommendations. The study was approved by the local IRBs at the Rabin, Shaare Zedek, Emek and Soroka Medical Centers, (1072-20-RMC, 0557-20-SZMC, 0247-20-EMC, 0568-20-SOR, respectively). MOH number: 2020-12-30_009617. All participants signed an informed consent form before any study procedure.

### Study procedure

Eligible participants were evaluated at 4-time points: (i) before the first vaccine dose -V1, (ii) 14-21 days after the first and before the second vaccine dose – V2, (iii) phone call a week after the second vaccine dose to report adverse events (AEs) and (iv) 21-35 days after the second vaccine dose – V3, (see Figure 1A). At enrolment, patients were assessed for baseline demographic and IBD characteristics. Specifically, medical treatment, duration and dose were registered, including date of biologics injections/infusions as well as interval between biologics administration and vaccination. Each visit clinical evaluation was performed using IBD specific questionnaires – Harvey-Bradshaw Index (HBI)^19^, Simple Clinical Colitis Activity Index (SCCAI)^20^ and Pouch Disease Activity Index (PDAI)^21^ for CD, UC and patients with an ileal pouch, respectively. Post-vaccination AEs^22^ were evaluated by standard questionnaires, specifically referring to pain or swelling at injection site, fever, headache, shivering, nausea, dizziness, fatigue, muscle soreness, joints pain, allergic reaction, other AEs^2, 22^, and severe AEs (SAEs, anaphylactic reaction, hospitalization, death). Safety measures also included assessment of IBD clinical activity as well as inflammatory biomarkers.

**Figure 1:**
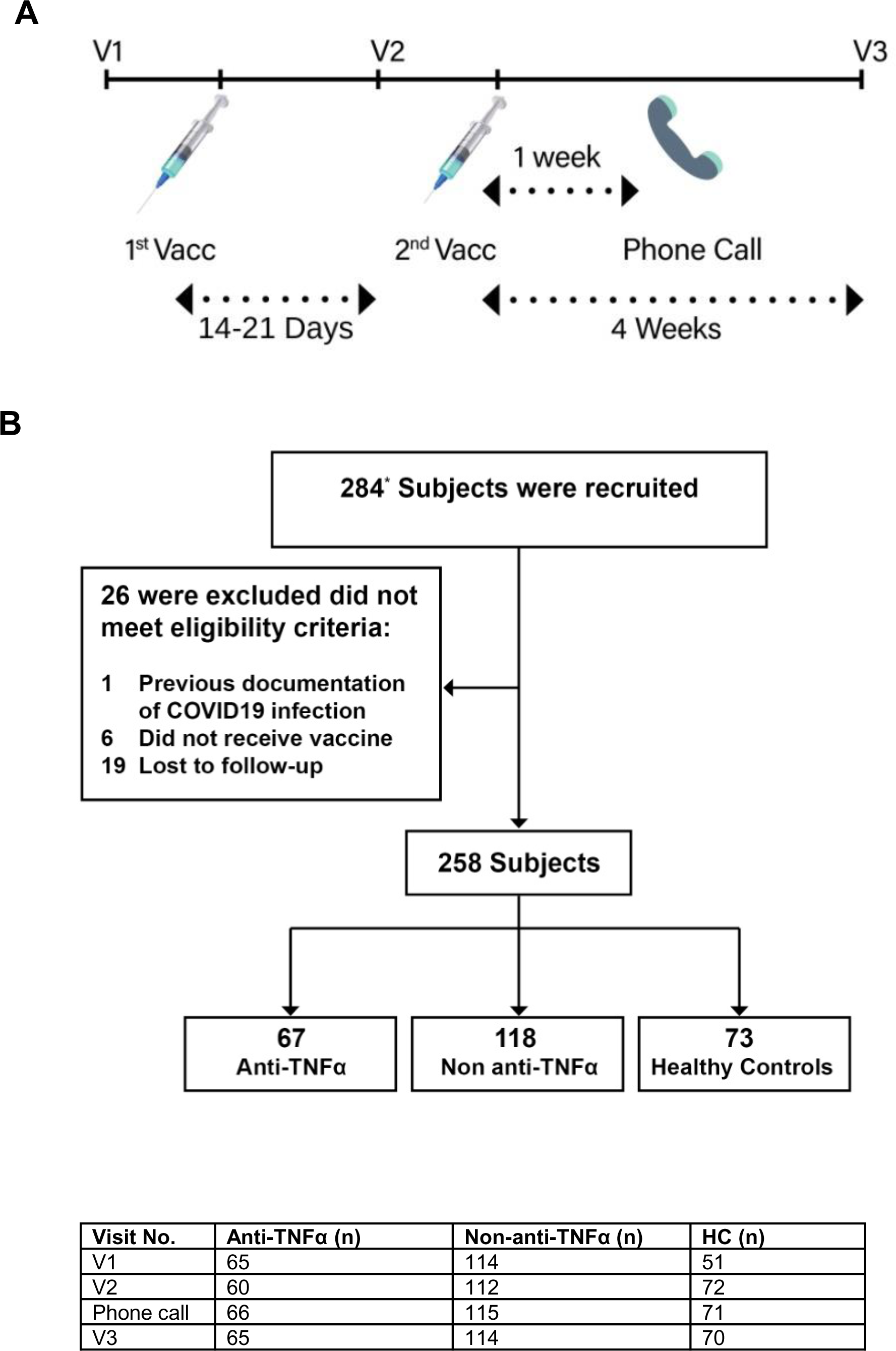
A. **Study protocol**. Patients were enrolled at visit 1 (V1), before the first vaccine dose. Second visit (V2) was 14-21 days after the first but before the second vaccine dose. A week after the second vaccine dose a phone call was made to evaluate adverse events (AEs), and a third visit (V3) was 4 weeks after the second vaccine dose. In each visit laboratory tests were performed, and questionnaires regarding disease severity and AEs were filled. B. **Patients disposition**. The diagram represents all enrolled participants who were recruited before vaccination. *28 subjects were recruited at the second visit (after first vaccine dose but before the second one), mainly due to logistic reasons. Most of them (22) were healthy controls (HC). Number of subjects at each visit is detailed in the table below the diagram. Abbreviations: HC=healthy controls, Vacc=vaccine dose.

Laboratory tests were performed at V1, V2 and V3 including complete blood count, C-reactive protein (CRP), COVID-19 serology and functional neutralization and inhibition assays. Anti-TNFα drug levels and anti-TNFα antibodies were measured. Serum was separated from collected blood, aliquoted and stored at -80°C until further analyses.

### Outcomes

The primary endpoint was seropositivity rate and magnitude of the immune response (levels of binding IgG antibodies to SARS-CoV-2 spike (S) antigen and neutralizing and inhibitory antibodies functionality) following BNT162b2 in patients with IBD with or without anti-TNFα treatment, or HC, at V3. Secondary endpoints were immune response dynamics induced after the first and second vaccine doses; and AEs, specifically local and systemic reactions and IBD exacerbation.

### Laboratory methods

**SARS-CoV-2 IgG II quantitative testing** was performed using the Abbott architect i2000sr platform in accordance with manufacturer’s instructions^23^. Values ≥50 activity units (AU)/mL are considered positive.

**SARS-CoV-2 nucleocapsid (N) IgG testing** was performed semiquantitatively using ELISA plates coated with N protein in accordance with manufacturer’s instructions (EUROIMMUN, Lubeck, Germany). Values ≥1.1 units are considered positive.

**Anti-TNFα drug and anti-drug antibody levels** were assessed for adalimumab (ADA and ADA-Abs) and infliximab (IFX and IFX-Abs) using Lisa-Tracker ELISA in accordance with manufacturer’s instructions (Theradiag, Beaubourg, France). Range for drug levels: 0.3-20 µg/mL. Range for Abs levels: 10-160 ng/mL and 10-200 ng/mL for ADA-Abs and IFX-Abs, respectively.

**Receptor binding domain (RBD): angiotensin converting enzyme (ACE)2 inhibition ELISA**^24^ was performed as described^25^ using RBD-serum mix incubated with ACE2 coated plates. Inhibition percentage was calculated for each well by the formula: 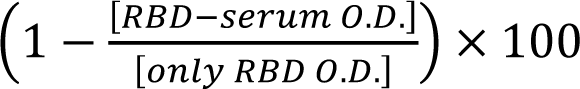. Negative results, indicating no inhibition, were set as 0% inhibition.

### Preparation of SARS-CoV-2-spike pseudoparticles and neutralization assay

To generate SARS-CoV-2 pseudo typed vesicular stomatitis virus (VSV) particles, human embryonic kidney (HEK)-293T cells were grown to 70% confluence in Dulbecco’s modified eagle medium (DMEM) supplemented in 10% fetal bovine serum (FBS), 1% L-glutamine, and 1% penicillin streptavidin. Cells were transfected with pCMV3 plasmid encoding the SARS-CoV-2 S protein with C-terminal, 19 residues truncation (pCMV3-SARS-CoV-2-SΔ19) using polyethylenimine (PEI). Twenty-four hours post-transfection, the cells were infected with G-complemented VSV^GFP^ΔG (*G-VSV^GFP^ΔG) at a multiplicity of infection (MOI) of 3. Following 6 hours incubation to allow internalization, cells were extensively washed 4 times with fresh medium to eliminate excess of *G-VSV^GFP^ΔG. After additional 30 hours of incubation the culture’s supernatant containing pseudotyped VSV (SΔ19-VSV^GFP^ΔG) was centrifuged (300×g, 5 min, 4 °C) to avoid cell debris, filtered on 0.2 µm filter cup, and stored in 1 mL aliquots at −80 °C until use. Titers were between 0.5×10^6^ to 1.5×10^6^ pseudovirus/mL.

HEK-293 cells stably expressing human ACE2 were cultured in DMEM (Biological Industries, Beit Haemek, Israel) supplemented in 10% FBS, 1% L-glutamine, and 1% penicillin streptavidin. These cells were seeded into 100 μg/mL poly-D-lysine-coated 96-well plates (Greiner-Bio-one, Kremsmünster, Austria) at an initial density of 0.5×10^5^ cells per well. The following day concentrated pseudo-particles were incubated with sera samples at dilution of 1:200 for 1 hour at 37°C and then added to the 96-well pre-seeded plates. After 24 h, medium was replaced with fresh DMEM excluding phenol red and plates were imaged by the IncuCyte ZOOM system (Essen BioScience, Michigan, USA). Cells were imaged with a 10X objective using the default IncuCyte software settings, which were used to calculate number of GFP-positive cells from four 488 nm-channel images in each well (data were collected in triplicate). The number of GFP-positive cells was normalized and converted to a neutralization percentage in each sample, compared to the average of control samples.

### Statistical Analysis

Data were collected in secured web-based platform (REDCap) and analyzed using SPSS version 27 (IBM, New York, United States).

All tests were two tailed and p<0.05 was considered significant. Anti-S antibody concentrations are expressed as geometric mean concentrations (GMCs) with 95% confidence intervals (CI). Other continuous data are reported as median and IQR unless otherwise stated. Counts and percentages were employed for categorical variables. Univariate analyses, using independent samples t-test, one-way analysis of variance (ANOVA) with Bonferroni multiple-comparison correction or Kruskal-Wallis nonparametric test of ln-transformed anti-S antibody concentration and Spearman’s rank correlation coefficients, were used to identify demographic, disease, vaccine, and treatment-related factors associated with anti-S levels. We used multivariate stepwise linear regression models to identify factors independently associated with ln anti-S levels. Standardized Beta coefficients were obtained from linear regression.

## Results

### Study population

Subjects were recruited in IBD centers located in central (Rabin); Northern (Emek); Eastern and Jerusalem (Shaare Zedek); and Southern (Soroka) Israel, between December 29^th^, 2020, and May 5^th^, 2021. Participants’ baseline characteristics are presented in Table 1. Of 258 subjects, 185 had IBD (122 CD, 53 UC, 6 IPAA, 4 IBD-Unclassified [U]) and 73 were HC. Average age (years) in the IBD (37.9±14.3) and HC groups (36.6±12.4) was comparable. There were 60.6% males in the IBD and 27.4% in the HC group. The majority (56/67) of patients treated with anti-TNFα had CD. Patient disposition is presented in figure 1B. In the anti-TNFα group concomitant therapy included: immunomodulators (8), 5-ASA (5) and steroids (1). Concomitant therapy in the non-anti-TNFα group included: 5-ASA (37), non-anti-TNFα biologics (34, mostly vedolizumab), immunomodulators (8), steroids (7), and tofacitinib (3). There were 38 untreated patients (26 CD, 8 UC, 4 IPAA). Most subjects (230/258, 89%) were recruited within a median of 1 (Interquartile range [IQR] 0-4) days before the first vaccine dose (V1), and 28/258 (10.8%) within 21 (IQR 20-21) days post first vaccine dose (V2) mainly due to logistic reasons (Figure 1B). Median interval between first and second vaccine doses was 21(IQR 20-24) days. Median interval between the second vaccine dose and V3 blood sampling was 30 (IQR 28-33) days. Baseline laboratory results, including blood counts and CRP were comparable between the groups (Supplementary Table 1).

**Table 1:** Baseline demographic characteristics of participants

| Characteristic | Anti-TNFα<br>N=67 | Non-anti-TNFα<br>N=118 | HC<br>N=73 | P value |
| --- | --- | --- | --- | --- |
| Mean age, years (SD) | 37.8 (14.3) | 38.2 (14.3) | 36.6 (12.4) | 0.744 |
| Female, n (%) | 24 (35.8) | 49 (41.5) | 53 (72.6) | **<0.001** |
| Origin, n (%) |  |  |  |  |
| Ashkenazi | 31 (46.3) | 49 (41.5) | 36 (49.3) | 0.558 |
| Non-Ashkenazi | 36 (53.7) | 69 (58.5) | 37 (50.7) |  |
| Mean BMI, kg/m<sup>2</sup> (SD) | 25 (4.0) | 24.4 (5.2) | 25.7 (6.4) | 0.354 |
| Smoking status, n (%) |  |  |  |  |
| Present | 8 (11.9) | 15 (12.7) | 7 (9.5) | 0.299 |
| Past | 9 (13.4) | 7 (5.9) | 3 (4.1) |  |
| No | 50 (74.6) | 89 (75.4) | 63 (86.3) |  |
| Comorbidities<sup>a</sup>, n (%) | 8 (11.9) | 11 (9.3) | 5 (6.8) |  |
| IBD phenotype, n (%) |  |  |  |  |
| CD | 56 (83.6) | 66 (55.9) | ----- | **<0.001** |
| UC | 8 (11.9) | 45 (38.1) | ----- | **<0.001** |
| IPAA | 2 (3) | 4 (3.4) | ----- |  |
| IBD-U | 1 (1.5) | 3 (2.5) | ----- |  |
| Disease activity<sup>b</sup>, n (%) |  |  |  |  |
| Remission | 46 (68.6) | 74 (62.7) | ----- | 1.000 |
| Active | 21 (31.4) | 44 (37.3) | ----- |  |
| Current medication, n (%) |  |  |  |  |
| IFX | 34 (50.7) | ----- | ----- |  |
| ADA | 33 (49.3) | ----- | ----- |  |
| Vedolizumab | ----- | 26 (22.03) | ----- |  |
| Ustekinumab | ----- | 5 (4.23) | ----- |  |
| 5-ASA | 5 (7.4) | 37 (31.3) | ----- |  |
| Steroids | 1 (1.5) | 7 (5.9) | ----- |  |
| Immunomodulators<sup>c</sup> | 8 (11.9) | 8 (6.7) | ----- |  |
| JAK inhibitor | ----- | 3 (2.5) | ----- |  |
| No treatment | ----- | 38 (32.2) | ----- |  |

<sup>a</sup>Comorbidities were present in 21 patients overall and included mainly asthma (6), diabetes (5), high blood pressure (5) and celiac (2). The rest were fatty liver disease, hypothyroidism, ankylosing spondylitis, and prostate cancer.
<sup>b</sup>Disease activity was quantified clinically by validated questionnaires.
<sup>c</sup>Including 6-mercaptopurine, azathioprine, methotrexate.
Abbreviations: HC=healthy controls, BMI=body mass index, CD=Crohn's disease, UC=ulcerative colitis, IBD-U=IBD-unclassified, IPAA=ileal pouch anal-anastomosis, IFX=infliximab, ADA=adalimumab, 5-ASA= 5-aminosalicylic acid.

### All patients with IBD achieve seropositivity after the second vaccine dose, however those treated with anti-TNFα have significantly lower antibody titers

SARS-CoV-2 anti-S IgG antibodies were positive in all subjects after the second vaccine dose (V3). This suggests that neither IBD itself nor anti-TNFα treatment abolish the ability to mount an immune response to two BNT162b2 doses. However, anti-TNFα treatment was associated with significantly lower antibody levels. Specifically, pre-vaccination (V1), anti-S IgG GMCs were negligible in all subjects (Figure 2A; Supplementary Table 2). V2 GMCs (95% CI) were 2-3-fold lower in the anti-TNFα treated compared to untreated patients and HC groups: 340 (221-523), 710 (509-991), and 1039 (797-1355), p=0.012 and p<0.001, respectively (Figure 2B). GMC increase after the second vaccine dose was robust and similar (around 10-fold) in all study groups maintaining the 2-3-fold differences between the groups. V3 GMCs (95% CI) were 3787 (2732-5249), 8320 (6630-10441), and 10979 (9396-12829), p<0.001 and p<0.001, in the anti-TNFα treated compared to the untreated patients and HC groups, respectively (Figure 2C). Importantly, while all HC were seropositive at V2 (no subject< 50 AU/mL), 14 patients with IBD were still seronegative, of whom 6 were treated and 8 untreated with anti-TNFα (Figure 2D).

**Figure 2:**
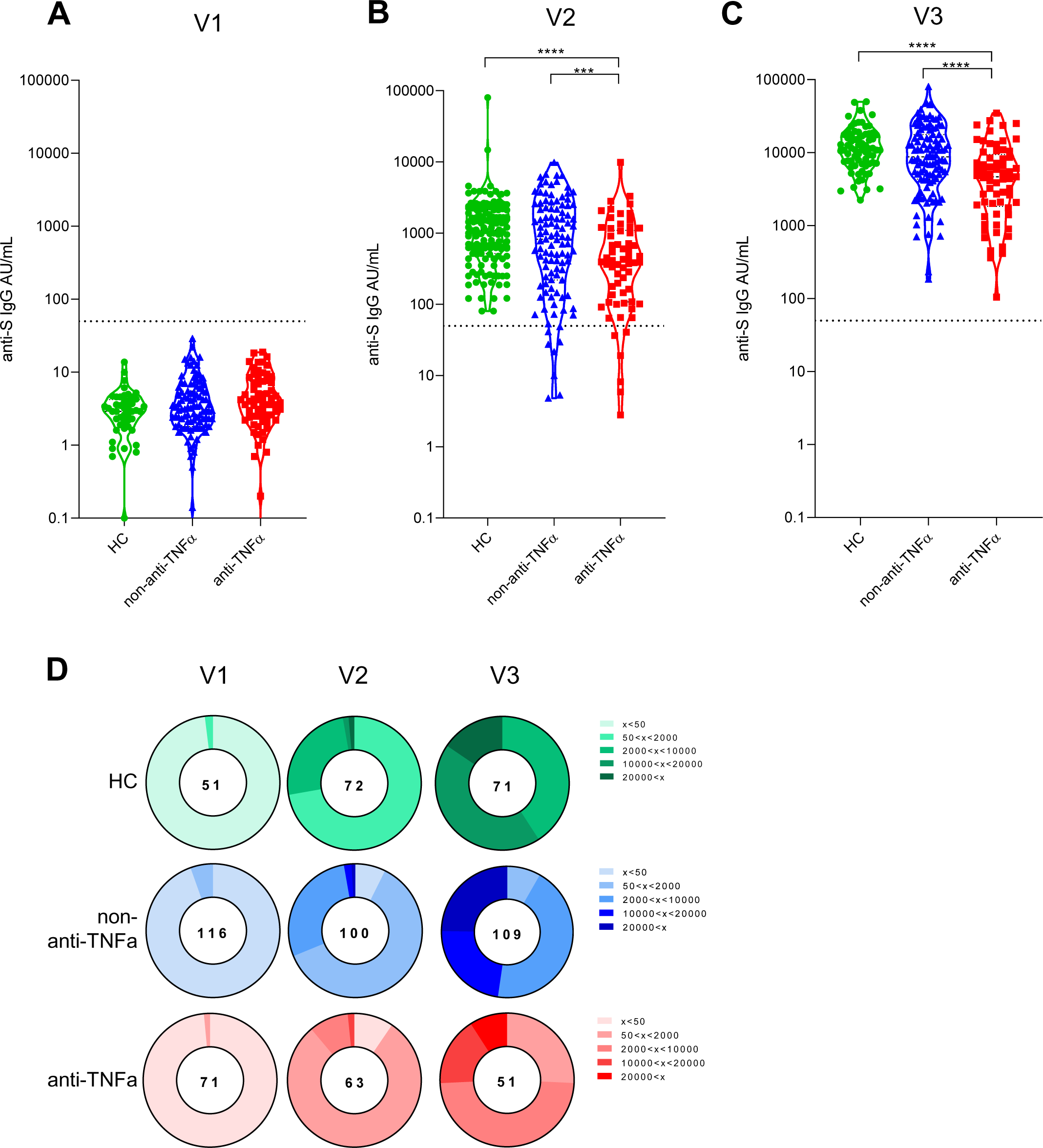
Patients with IBD treated with anti-TNFα have significantly reduced levels of anti-S antibodies. (A-C) Levels of anti-S antibodies in sera from healthy controls (HC, shown in green), patients with IBD receiving non-anti-TNFα treatment (non-anti-TNFα, shown in blue) and patients with IBD receiving anti-TNFα treatment (anti-TNFα, shown in red). Antibodies were measured by the Abbott quantitative anti-S IgG kit. Visit 1 (V1) – before vaccination, visit 2 (V2) and visit 3 (V3), after first and second vaccine doses, respectively. Statistical analysis was carried out using independent-samples Kruskal-Wallis test *** - p< 0.0005, **** - p< 0.0001 (D) Pie charts representing the fractions of patients at timepoints V1, V2 and V3, with anti-SARS-CoV-2 antibodies levels as designated in the legend (A-C). Numbers in the middle of the pies denote the total number of subjects tested in each group for every timepoint.

We next assessed neutralizing antibodies, considered critical for patients survival and virus control^26^. Using competitive ELISA we show that while at V1 inhibition activity was low and comparable between the groups (Figure 3, A-C, Supplementary Table 3), at V2 the anti-TNFα treated group had significantly lower ability to inhibit RBD:ACE2 binding compared to HC (p<0.05). This was even more prominent at V3 (p<0.001). Notably, significant differences in inhibition activity were apparent between patients with IBD, regardless of treatment regimen, and HC (Figure 3C). We observed a positive correlation between anti-S titers and inhibition activity in V2, which increased even further in V3, suggesting that after two vaccine doses the proportion of anti-S IgG antibodies with inhibitory function increases (Figure 3, E-F).

**Figure 3:**
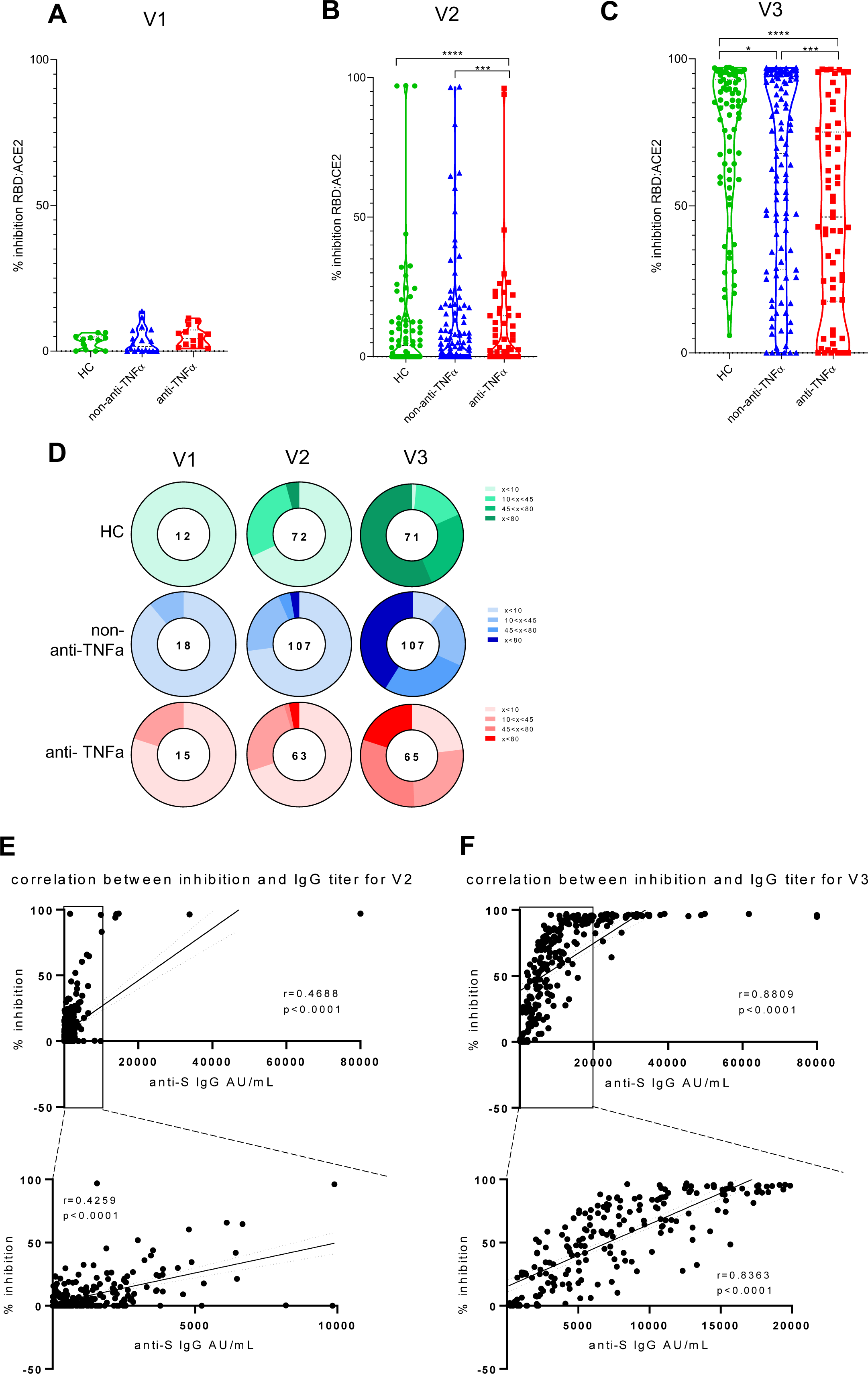
Patients with IBD treated with anti-TNFα have significantly reduced levels of anti-SARS-CoV-2 inhibiting antibodies. (A-C) Ability of serum from healthy controls (HC, shown in green), patients with IBD receiving non-anti-TNFα treatment (non-anti-TNFα, shown in blue) and patients with IBD receiving anti-TNFα treatment (anti-TNFα, shown in red) to inhibit SARS-CoV-2 RBD binding to ACE2 receptor. Values measured by ELISA are presented as % inhibition (y axis), following vaccination. Visit 1 (V1) – before vaccination, visit 2 (V2) and visit 3 (V3), after first and second vaccine doses, respectively. Zero inhibition was set as the value of RBD without added sera. Statistical analysis was carried out using independent-samples Kruskal-Wallis test *** - p< 0.0005, **** - p<0.0001. At least three repetitions for every sample. (D) Pie charts representing the fractions of patients at timepoints V1, V2 and V3, who developed none (<20%), low (20%<x<50%), medium (50%<x<80%), and high (>80%) SARS-CoV-2 RBD:ACE2 inhibition, based on (A-C). The numbers in the middle of the pies denote the total number of subjects tested in each group for every timepoint. (E-F) Graphs show correlation between anti-S titer measured in figure 2 and sera inhibition for all subjects. Left and right panels represent V2 and V3 timepoints, respectively. The lower panels are zoom-in view for each of the black frame-surrounded portions from the upper panel. Correlation was calculated by Pearson correlation analysis. Abbreviations: RBD= receptor-binding domain, ACE2= angiotensin converting enzyme-2

Finally, we assessed vaccine functional activity using SARS-CoV-2 spike pseudoparticles neutralization assays. Serum from patients in all groups did not neutralize infection in V1, and was used for normalizing neutralization at V2 and V3. At V2 HC serum had a 65% neutralization capability, contrasting with significantly reduced activity in the anti-TNFα group (51%, p<0.05; Supplementary Table 4). Furthermore, at V3 serum from the HC and the non-anti-TNFα treated groups had significantly higher neutralization activity compared to serum from patients in the anti-TNFα group (97%, 96%, and 79%, respectively, p<0.0001; Figure 4, A, B). Neutralization activity highly correlated with both anti-S titers and inhibition measures (Figure 4,D-F), suggesting that anti-S IgG assays may be indicative of the functional serological anti-SARS-CoV-2 antibody activity.

**Figure 4:**
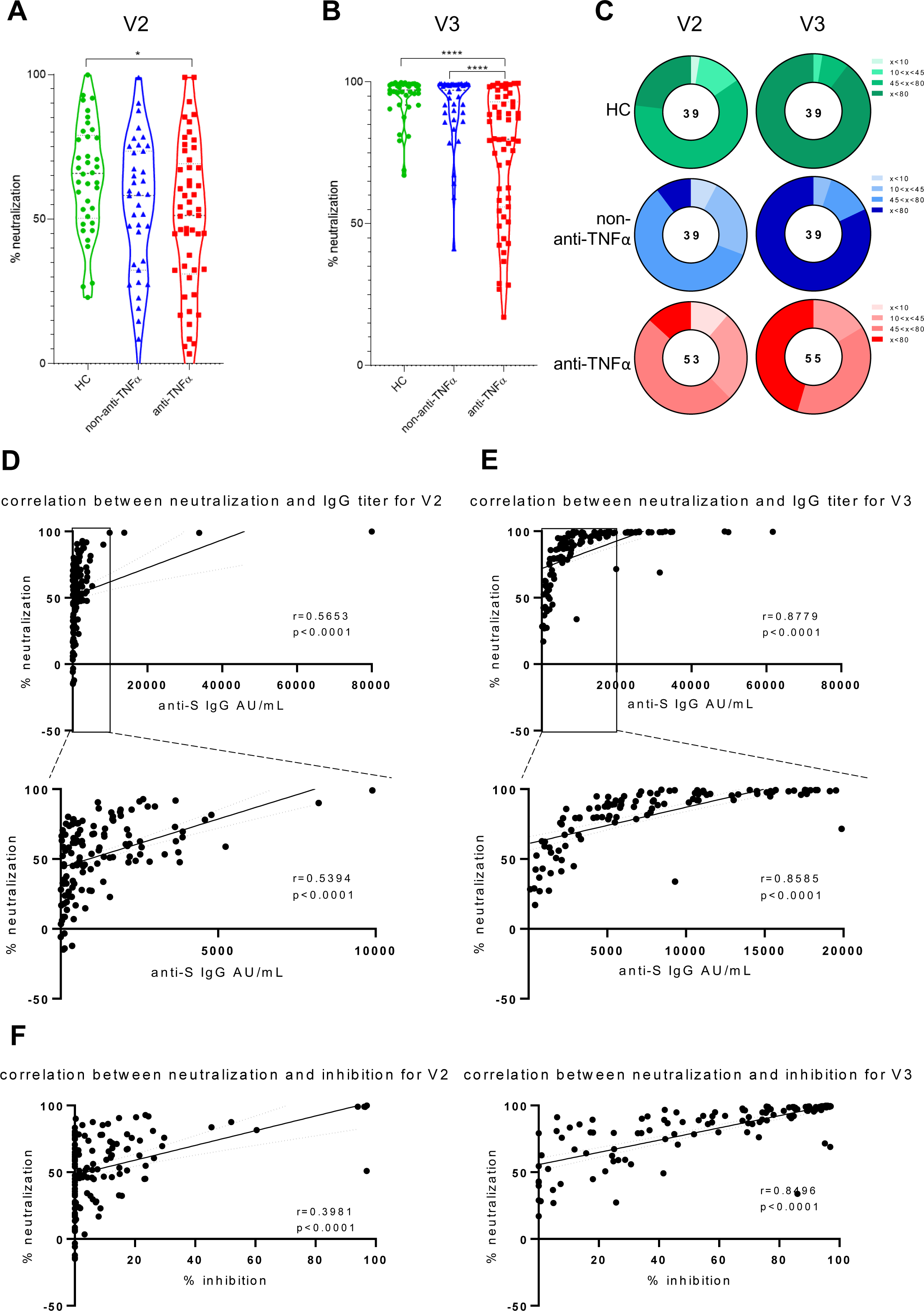
Patients with IBD treated with anti-TNFα have significantly reduced levels of anti-SARS-CoV-2 neutralizing antibodies. (A, B) Sera, diluted to a final concentration of 1:200, were incubated with VSV-spike pseudo-particles (VSVΔG^GFP^SΔ19) for 1 hour in 37°C, prior to infecting ACE2 expressing HEK293 cells for 24 hours. The number of GFP-positive cells was normalized and converted to a neutralization percentage in each sample, compared to the average of control samples. Visit 1 (V1) – before vaccination, visit 2 (V2) and visit 3 (V3), after first and second vaccine doses, respectively. Statistical analysis was carried out using independent-samples Kruskal-Wallis test *** - p< 0.0005, **** - p< 0.0001 (C) Pie charts representing the fractions of patients in timepoints V2 and V3, who developed none (<20%), low (20%<x<50%), medium (50%<x<80%), and high (>80%) SARS-CoV-2 RBD neutralizing antibodies, based on (A, B). (D-E) Graphs show correlation between anti-S titer measured in figure 2 and sera neutralization for all donors. Left and right panels represent V2 and V3 timepoints, respectively. The lower panels are zoom-in view for each of the black frame-surrounded portions from the upper panel. (F) Graphs show correlation between sera inhibition measured in figure 3 and sera neutralization for all donors. Left and right panels represent V2 and V3 timepoints, respectively. All correlations were calculated by Pearson correlation analysis. Abbreviations: VSV= vesicular stomatitis virus, ACE2= angiotensin converting enzyme-2, RBD= receptor-binding domain, HEK= human embryonic kidney.

Importantly, non-anti-TNFα IBD therapies did not significantly modify seroconversion or magnitude of response. Specifically, in patients treated with vedolizumab (n=27, 51.8% UC) or 5-ASA (n=35, 76.9% UC), vaccine responses were comparable to those of HC (p=0.288, 0.191, respectively).

Anti-N, reflecting infection with COVID-19 was positive after the second vaccine dose in <2% of study participants and comparable between the groups. Specifically, anti-N Abs were detected in 2 HC, 2 non-anti-TNFα and 1 anti-TNFα treated patients. These subjects were not excluded from analysis given the equal distribution between the groups and the comparable to not-infected patients anti-S titers.

### Older age is an additional predictor of lower vaccine immune response

In univariate analysis (Supplementary Table 5) factors such as older age, male gender and WBC were also associated with a lower serologic response after the first vaccine dose (male gender and WBC value) and after both vaccine doses (older age).

In multivariate linear regression model only anti-TNFα treatment and older age maintained a significant distinct association with lower IgG anti-S response (Supplementary Table 6).

The inverse correlation between older age and lower IgG anti-S antibodies levels in the three study groups after the first and second vaccine doses is displayed in Supplementary Figure 1, while Supplementary Table 7 shows the consistently lower GMCs in subjects 40 years and above compared with younger ones in all study groups after the first and second vaccine doses.

### Anti-TNFα drug levels at the time of vaccination do not affect immune responses

We next asked whether anti-TNFα drug levels mediated lower vaccine immune responses in this group. Importantly, anti-TNFα drug level measurement was not assessed at trough (i.e., immediately prior to anti-TNFα drug administration), but at the time of serologic assessment at V1, V2 and V3. No correlation between drug levels and immune responses was observed (Supplementary Table 8). We further asked whether lower responses in patients treated with anti-TNFα were affected by the interval between anti-TNFα drug administration and vaccination. Importantly, no such correlation was observed neither when anti-TNFα drugs were administered before the first or second vaccine doses (Supplementary Figure 2). Finally, only two patients had anti-IFX and two anti-ADA drug antibodies. Those did not correlate with vaccine immune responses (Supplementary Table 9).

### Vaccine is safe in patients with IBD and is not associated with increased IBD activity

Immediate and short-term AEs were detected using phone call and accepted questionnaires, respectively. We further evaluated IBD exacerbation using clinical and laboratory variables. To this end, no SAEs were registered. The most common AEs were local pain and headache, with more AEs after the second compared to first vaccine dose (Supplementary Table 10). AEs were not in excess or more prominent in patients treated with anti-TNFα who had higher drug levels during vaccination.

Finally, baseline IBD activity was comparable in patients treated with anti-TNFα or not and remained comparable after the first and second vaccine doses (Supplementary Table 11, Supplementary Figure 3). Neither CRP levels nor WBC count were increased following vaccination in both groups.

## Discussion

Patients with IBD treated with immunomodulators and/or anti-TNFα biologics are at an increased risk of vaccine preventable diseases, and vaccination programs are recommended. Patients with chronic diseases were more prone to COVID-19 complications and death^27–29^. Vaccination campaigns encouraged patients with IBD to vaccinate^30, 31^, despite their exclusion from phase-3 trials^2, 3^. Here, we aimed to prospectively evaluate immune responses and safety of the BNT162b2 vaccine in patients with IBD.

Our results show that all subjects, regardless of medical treatment, seroconverted after the second vaccine dose. However, patients treated with anti-TNFα had significantly lower immune responses, represented by 2-3-fold decreased IgG anti-S levels, compared to patients untreated with anti-TNFα and HC. Furthermore, impaired immune function was demonstrated by significantly lower RBD:ACE2 inhibition and significantly lower capability to neutralize SARS-CoV-2 in a pseudoviral assay.

According to a recent population-based report, patients with IBD treated with anti-TNFα had a lower serologic response to COVID-19 infection^32^ and vaccination^33^, in line with previous reports regarding other vaccines^10, 13, 15–17, 33–35^. Our study is the first to prospectively and comprehensively demonstrate the profound impairment in functional immune responses in anti-TNFα treated patients with IBD, which may be relevant for other immune mediated diseases as well^25^. Reassuringly, the rate of anti-N Abs was low and comparable in all groups suggesting that protection was enough to prevent short-term infection. However, our data also suggest that the duration of protection may be shorter in patients treated with anti-TNFα compared to non-anti-TNFα treated patients or HC. If indeed both longevity and neutralizing activity of anti SARS-CoV-2 antibodies is reduced, the consequence may be reduction in infection protection, supporting earlier booster vaccination for this patient subpopulation, as already considered for severely immunocompromised patients such as those with certain cancers and chemotherapy ^36, 37^.

The prospective nature of our study enabled evaluation of immune responses dynamics. Importantly, about 10% of patients treated with anti-TNFα were still seronegative after the first vaccine dose, and additional 18% had a low level (50-150 AU) of anti-S antibodies. This supports maintainance of thorough COVID-19 precautions for them and their household members until after the second vaccine dose. Notably, after the first vaccine dose there were also 8 (7%) seronegative patients in the non-anti-TNFα treated group pointing to additional patient factors that may modify seronegativity.

In this regard, we found that age was an independent predictor of lower vaccine immune responses, regardless of IBD treatment. While our patients were mostly young (∼37 years), a continuous decline in serology with age was noticed. As older age is also a risk factor for severe COVID-19^38, 39^ these patients should be at highest priority for booster vaccine doses. A recent report from US Veteran Affairs data base demonstrating only 80.4% vaccine effectiveness in a patient population with a median age of 68 supports our finding^40^.

Our study, the first specifically designed to adress vaccine timing relative to anti-TNFα drug administration did not demonstrate such correlation. Moreover, in 14 patients vaccinated during anti-TNFα induction, responses were comparable to those vaccinated during maintenance. Anti-TNFα drug levels during vaccination, were unrelated to impaired responses. Altogether, these findings lend evidence to the empiric recommendation to vaccinate patients with IBD regardless of anti-TNFα administration timing^30^.

Importantly, no SAEs were reported in the week following vaccination. While our study included only 10 subjects< 21 years, it is reassuring that no cardiac AEs, specifically myocarditis^38^ were detected. AEs were similar in all subjects-mainly local pain, and resolved within a few days. No IBD exacerbation was observed, regardless of disease activity during vaccination. This is specifically reassuring as approximately a third of patients were not in remission.

There are several strengths to our study. This is the first prospective multi-center study comprehensively investigating multiple aspects of BNT162b2 vaccine immune responses in patients with IBD. Most subjects were recruited before the first vaccine dose, allowing longitudinal evaluation of the dynamics of immune response development. While focusing on anti-TNFα therapy, all other IBD therapies, or no therapy were included. Thus, enabling differentiation between disease and treatment effects. Finally, this is the first study addressing timing of vaccination and anti-TNFα drug administration, levels, or anti-drug antibodies, showing a lack of correlation. Another meaningful strength is assessment of vaccine safety including IBD activity, as previous reports in other immune mediated diseases suggested disease exacerbation post-vaccination^41–46^.

Our study, including 67 patients treated with anti-TNFα was powered to demonstrate significant differences, which indeed were apparent, between them and untreated patients. Limitations include difference in gender ratio between IBD and HC groups at baseline, the relatively young age of participants (although this reflects typical IBD populations) and the use of only one vaccine type. Evaluation of vaccine efficacy is limited, as infection rate in Israel during the study period was low. Finally, observation was limited to 4 weeks after the second vaccine.

To conclude, our study provides prospective, controlled evidence for the efficacy and safety of the COVID-19 BNT162b2 vaccine in patients with IBD stratified according to therapy. We demonstrate the dynamics of development of functional immune response and the factors causing impairment, specifically anti-TNFα therapy and older age. The lack of correlation with timing of anti-TNFα therapy or drug levels, enables important clinical guidance to patients and their caregivers.

As immune response longevity in this group may be limited, vaccine booster dose should be considered.

Long(er) term outcomes and the mechanism of decreased immune responses should be evaluated.

## Data Availability

no data available

## List of abbreviations

5-ASA: 5-aminosalicylic acid
Ab: antibodies
ACE2: angiotensin converting enzyme2
ADA: adalimumab
AEs: adverse events
Anti-TNFα: anti-tumor necrosis factor (TNF) α
AU: activity units
BMI: body mass index
CI: confidence intervals
COVID-19: Corona Virus 2019
CRP: C-reactive protein
GMC: as geometric mean concentration
HBI: Harvey-Bradshaw Index
HC: healthy controls
HEK: human embryonic kidney
IBD-U: IBD-Unclassified
IFX: infliximab
IPAA: ileal pouch anal-anastomosis
IQR: Interquartile range
N: nucleocapsid
PDAI: Pouch Disease Activity Index
RBD: Receptor binding domain
S: spike
SAEs: severe adverse events
SARS-CoV-2: Severe acute respiratory syndrome coronavirus 2
SCCAI: Simple Clinical Colitis Activity Index
VSV: vesicular stomatitis virus

## Disclosures

### ID: Consultation/advisory board

Abbvie,Athos, Arena, Cambridge Healthcare, Celltrion, Celgene/BMS,DSM, Ferring, Food Industries Organization, Iterative Scopes,Integra Holdings, Janssen, MSD, Neopharm,Pfizer,Rafa laboratories, Roche/Genentech, Sangamo, Sublimity, Takeda,Wildbio

### Speaking/teaching

Abbvie, Altman, Celltrion, Celgene/BMS, Ferring, Falk Pharma, Janssen, MSD, Neopharm, Nestle, Pfizer, Rafa laboratories, Roche/Genentech, Sandoz, Takeda

### ABGS

Grant support from Takeda and Janssen, consultancy and lectures fees from Takeda, Janssen, Abbvie, Pfizer, Neopharm and BMS.

EZ: has received research support and consulting fees from Janssen, Abbvie, Takeda,

Neopharm, Celgene and Pfizer.

All other authors declare they have no conflict of interest.

### Transcript Profiling

None

### Writing Assistnace

None

### Author contribution

study concept and design (ID, HEK, KMR, MHP, AF); acquisition of data (HEK, EZ, ABGS, IG, IAB, JEO, LL, HEB, HY, YS, AF, MHP, ALB, AS, YB, EM, BO, MAG, ES, SBH, TTP, RE, NK, MN, MW, JA); analysis and interpretation of data (ID, DC, NF, MGT, HEK, KMR, MN, MW, JA, MD); drafting of the manuscript (HEK, ID); critical revision of the manuscript for important intellectual content (ID, DC, NF, MGT); statistical analysis (SG, DC); obtained funding (NF, MGT, ID); administrative, technical, or material support (ID, DC, KMR, NF, MGT); study supervision (ID)

## Acknowledgements

We would like to thank study participants for their effort and time. The Israeli IBD Society and Israeli GI association, are thanked for their support, and Dr. Naim Abu-Freha for specific efforts to promote the study. The study was partially supported by a generous grant from The Leona M. and Harry B. Helmsley Charitable Trust. The Crohn’s and Colitis Foundation of Israel, the European Federation of Crohn’s and Colitis Associations and the AMICI group are thanked for partially funding the study and supporting recruitment efforts. We thank Kawsar Kaboub and Hanan Abu Taha from the Mucosal Immunology Laboratory at the Felsenstein Medical Research Center for their scientific contribution. This study was performed in collaboration with the Israeli Ministry of Health. NTF acknowledges the kind support of L. Cohen and the Milner Foundation.

## Supplementary Material

**Supplementary Table 1:** Selected laboratory tests at baseline.

| Test, mean (SD) | Anti-TNFα | Non-anti-TNFα | HC | P value |
| --- | --- | --- | --- | --- |
| Hemoglobin [g/dL] | 14 (1.4) | 13.6 (1.8) | 13.6 (1.3) | 0.144 |
| White blood cells [K/μL] | 7.2 (2.1) | 7.1 (2.1) | 7 (1.99) | 0.863 |
| CRP [mg/dL] | 0.6 (1.1) | 0.7 (1.1) | 0.4 (0.6) | 0.126 |

Values are mean (SD). Abbreviations: CRP=C-reactive protein, HC=healthy controls.

**Supplementary Table 2:** Geometric Mean Concentrations (GMCs) of IgG against the S-antigen of the 3 groups at each visit.

| Visit | Group | GMC | 95% Confidence Interval for GMC |  | ANOVA |  |  |
| --- | --- | --- | --- | --- | --- | --- | --- |
| | | | Lower Bound | Upper Bound | P value HC | P value Non-anti-TNFα | P value Anti-TNFα |
| V1 | HC | 3.2 | 2.2 | 4.7 | ----- | 0.403 | 0.879 |
| | Non-TNFα | 4.5 | 3.4 | 5.8 | 0.403 | ----- | 1.000 |
| | TNFα | 4.1 | 3.3 | 5.2 | 0.879 | 1.000 | ----- |
| V2 | HC | 1039.8 | 797.6 | 1355.5 | ----- | 0.333 | <0.001* |
| | Non-TNFα | 710.5 | 509.2 | 991.2 | 0.333 | ----- | <0.001* |
| | TNFα | 340.3 | 221.1 | 523.7 | <0.001* | <0.001* | ----- |
| V3 | HC | 10979.3 | 9396.0 | 12829.5 | ----- | 0.272 | <0.001* |
| | Non-TNFα | 8320.4 | 6630.0 | 10441.6 | 0.272 | ----- | <0.001* |
| | TNFα | 3787.0 | 2732.0 | 5249.3 | <0.001* | <0.001* | ----- |

This table emphasizes the significant difference in serologic response between anti-TNFα and non-anti-TNFα and HC after first and second vaccine doses.
\*Testing differences between groups with Bonferroni correction for multiple comparisons Abbreviations: HC=healthy controls.

**Supplementary Table 3:** Neutralizing antibodies, measured by percentage of inhibition by RBD:ACE2.

| Visit | Group | Median (IQR) |
| --- | --- | --- |
| V1 | Anti-TNFα | 5.19 (1.7-9.2) |
| | Non-anti-TNFα | 3.5 (0-7.12) |
|  | HC | 3.9 (0.1-4.9) |
| V2 | Anti-TNFα | 1.8 (0-14.6) |
| | Non-anti-TNFα | 2.36 (0-12.7) |
|  | HC | 3.99 (0-12.4) |
| V3 | Anti-TNFα | 44.5 (14.8-74.7) |
| | Non-anti-TNFα | 67.3 (28.1-93.9) |
|  | HC | 83.9 (58.9-92.9) |

Abbreviations: RBD= receptor-binding domain, ACE2= angiotensin converting enzyme-2, HC=healthy controls, IQR=interquartile range.

**Supplementary Table 4:** Pseudovirus inhibition.

| Visit | Group | Median (IQR) |
| --- | --- | --- |
| V2 | Anti-TNFα | 51.1 (30.3-69.8) |
| | Non-anti-TNFα | 58.2 (32.8-73.3) |
|  | HC | 65.5 (21.6-78.4) |
| V3 | Anti-TNFα | 79.8 (55.2-92.5) |
| | Non-anti-TNFα | 96.5 (85.9-99.1) |
|  | HC | 97.1 (91.8-99.2) |

Vaccine functional activity was assessed using SARS-CoV-2 spike pseudoparticles neutralization assays.
Abbreviations: HC=healthy controls, IQR= interquartile range.

**Supplementary Table 5:**
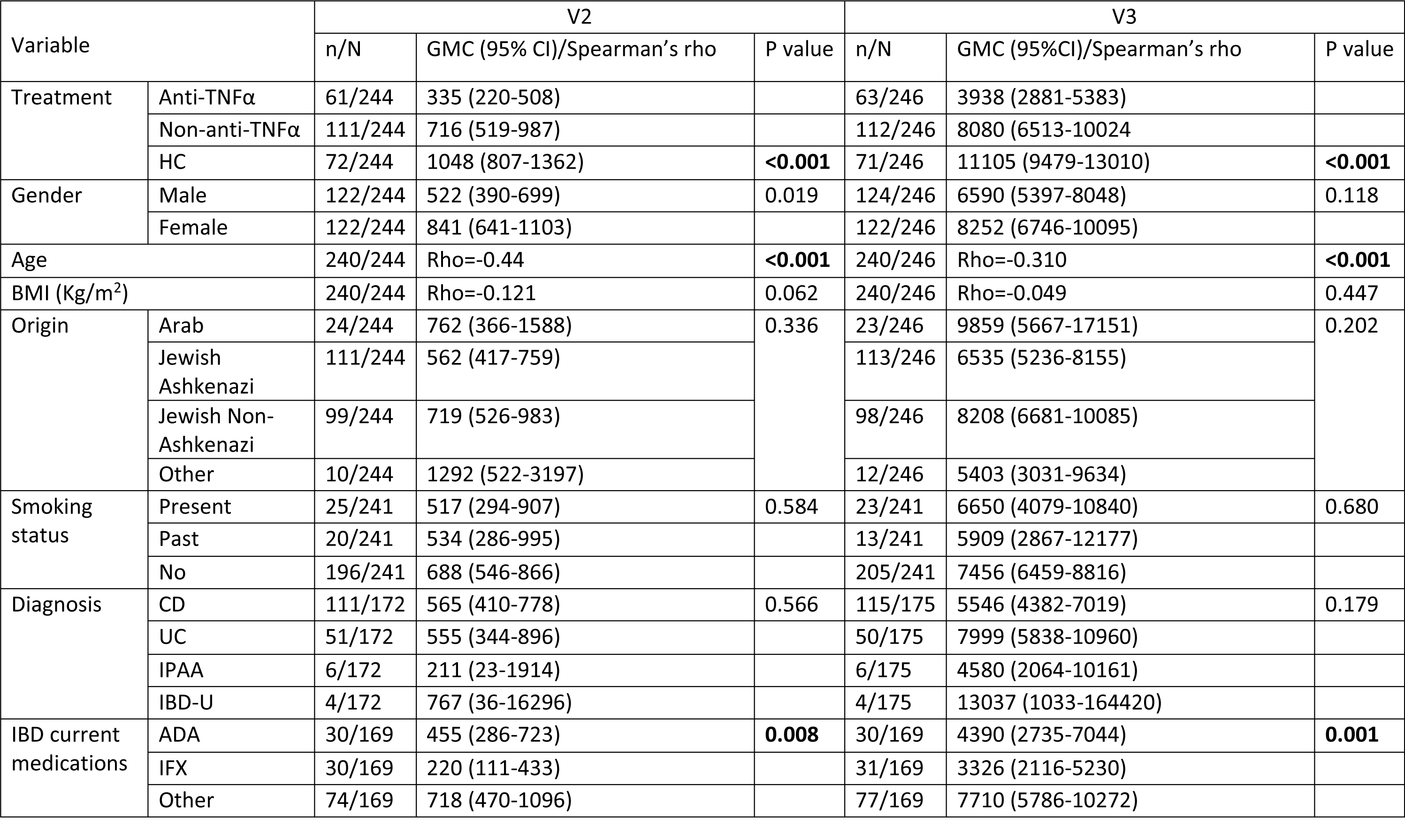
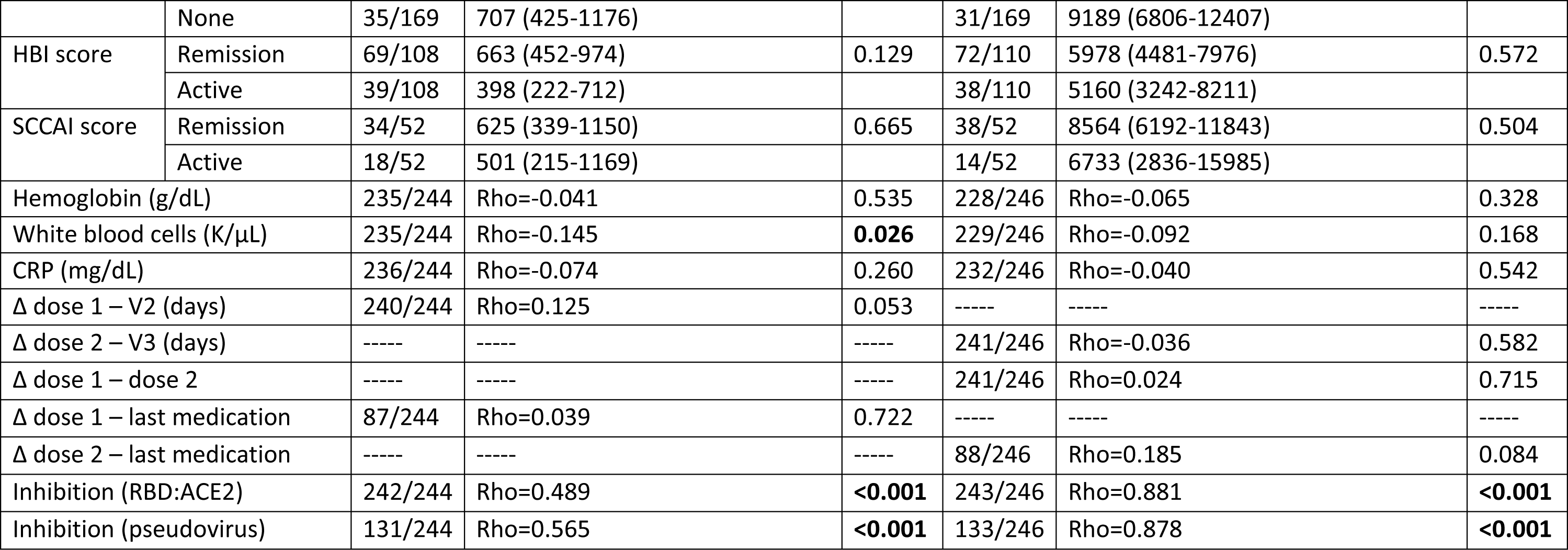
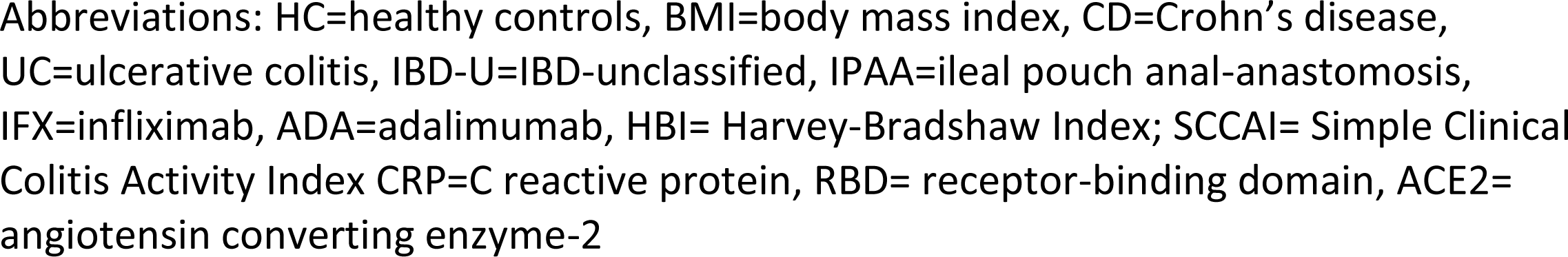
Factors associated with serologic response (univariate analysis)

| Variable |  | V2 |  |  | V3 |  |  |
| --- | --- | --- | --- | --- | --- | --- | --- |
|  |  | n/N | GMC (95% CI)/Spearman's rho | P value | n/N | GMC (95%CI)/Spearman's rho | P value |
| Treatment | Anti-TNFα | 61/244 | 335 (220-508) | | 63/246 | 3938 (2881-5383) | |
| | Non-anti-TNFα | 111/244 | 716 (519-987) | | 112/246 | 8080 (6513-10024) | |
|  | HC | 72/244 | 1048 (807-1362) | **<0.001** | 71/246 | 11105 (9479-13010) | **<0.001** |
| Gender | Male | 122/244 | 522 (390-699) | 0.019 | 124/246 | 6590 (5397-8048) | 0.118 |
|  | Female | 122/244 | 841 (641-1103) |  | 122/246 | 8252 (6746-10095) |  |
| Age |  | 240/244 | Rho=-0.44 | **<0.001** | 240/246 | Rho=-0.310 | **<0.001** |
| BMI (Kg/m<sup>2</sup>) |  | 240/244 | Rho=-0.121 | 0.062 | 240/246 | Rho=-0.049 | 0.447 |
| Origin | Arab | 24/244 | 762 (366-1588) | 0.336 | 23/246 | 9859 (5667-17151) | 0.202 |
|  | Jewish Ashkenazi | 111/244 | 562 (417-759) |  | 113/246 | 6535 (5236-8155) |  |
|  | Jewish Non-Ashkenazi | 99/244 | 719 (526-983) |  | 98/246 | 8208 (6681-10085) |  |
|  | Other | 10/244 | 1292 (522-3197) |  | 12/246 | 5403 (3031-9634) |  |
| Smoking status | Present | 25/241 | 517 (294-907) | 0.584 | 23/241 | 6650 (4079-10840) | 0.680 |
|  | Past | 20/241 | 534 (286-995) |  | 13/241 | 5909 (2867-12177) |  |
|  | No | 196/241 | 688 (546-866) |  | 205/241 | 7456 (6459-8816) |  |
| Diagnosis | CD | 111/172 | 565 (410-778) | 0.566 | 115/175 | 5546 (4382-7019) | 0.179 |
|  | UC | 51/172 | 555 (344-896) |  | 50/175 | 7999 (5838-10960) |  |
|  | IPAA | 6/172 | 211 (23-1914) |  | 6/175 | 4580 (2064-10161) |  |
|  | IBD-U | 4/172 | 767 (36-16296) |  | 4/175 | 13037 (1033-164420) |  |
| IBD current medications | ADA | 30/169 | 455 (286-723) | **0.008** | 30/169 | 4390 (2735-7044) | **0.001** |
|  | IFX | 30/169 | 220 (111-433) |  | 31/169 | 3326 (2116-5230) |  |
|  | Other | 74/169 | 718 (470-1096) |  | 77/169 | 7710 (5786-10272) |  |
|  | None | 35/169 | 707 (425-1176) |  | 31/169 | 9189 (6806-12407) |  |
| HBI score | Remission | 69/108 | 663 (452-974) | 0.129 | 72/110 | 5978 (4481-7976) | 0.572 |
|  | Active | 39/108 | 398 (222-712) |  | 38/110 | 5160 (3242-8211) |  |
| SCCAI score | Remission | 34/52 | 625 (339-1150) | 0.665 | 38/52 | 8564 (6192-11843) | 0.504 |
|  | Active | 18/52 | 501 (215-1169) |  | 14/52 | 6733 (2836-15985) |  |
| Hemoglobin (g/dL) |  | 235/244 | Rho=-0.041 | 0.535 | 228/246 | Rho=-0.065 | 0.328 |
| White blood cells (K/μL) | | 235/244 | Rho=-0.145 | **0.026** | 229/246 | Rho=-0.092 | 0.168 |
| CRP (mg/dL) |  | 236/244 | Rho=-0.074 | 0.260 | 232/246 | Rho=-0.040 | 0.542 |
| $\Delta$ dose 1 – V2 (days) | | 240/244 | Rho=0.125 | 0.053 | ----- | ----- | ----- |
| $\Delta$ dose 2 – V3 (days) | | ----- | ----- | ----- | 241/246 | Rho=-0.036 | 0.582 |
| $\Delta$ dose 1 – dose 2 | | ----- | ----- | ----- | 241/246 | Rho=0.024 | 0.715 |
| $\Delta$ dose 1 – last medication | | 87/244 | Rho=0.039 | 0.722 | ----- | ----- | ----- |
| $\Delta$ dose 2 – last medication | | ----- | ----- | ----- | 88/246 | Rho=0.185 | 0.084 |
| Inhibition (RBD:ACE2) |  | 242/244 | Rho=0.489 | **<0.001** | 243/246 | Rho=0.881 | **<0.001** |
| Inhibition (pseudovirus) |  | 131/244 | Rho=0.565 | **<0.001** | 133/246 | Rho=0.878 | **<0.001** |

Abbreviations: HC=healthy controls, BMI=body mass index, CD=Crohn's disease, UC=ulcerative colitis, IBD-U=IBD-unclassified, IPAA=ileal pouch anal-anastomosis, IFX=infliximab, ADA=adalimumab, HBI= Harvey-Bradshaw Index; SCCAI= Simple Clinical Colitis Activity Index CRP=C reactive protein, RBD= receptor-binding domain, ACE2= angiotensin converting enzyme-2

**Supplementary Table 6:** Factors associated with serologic response (multivariate linear regression). Standardized Beta coefficients were obtained from linear regression.

| Variable |  | V2 |  | V3 |  |
| --- | --- | --- | --- | --- | --- |
|  |  | B (95% CI) | P value | B (95% CI) | P value |
| Treatment | Anti-TNFα | -0.25 (-1.4-0.4) | **<0.001** | -0.32 (-1.2—0.46) | **<0.001** |
| | Non-anti-TNFα | -0.09 | 0.270 | -0.128 | 0.141 |
|  | HC | Reference |  | Reference |  |
| Gender | Male | -0.09 | 0.198 | -0.039 | 0.602 |
|  | Female | Reference |  | Reference |  |
| Age (years) |  | -0.43 (-0.07- -0.03) | **<0.001** | -0.27 (-0.03- -0.01) | **<0.001** |
| IBD current medication | ADA | 0.157 | 0.074 | 0.081 | 0.382 |
|  | IFX | -0.137 | 0.12 | -0.032 | 0.732 |
|  | Other | 0.007 | 0.935 | -0.114 | 0.262 |
|  | None | Reference |  | Reference |  |
| White blood cells (K/μL) | | -0.038 | 0.591 | -0.104 | 0.159 |

Abbreviations: HC=healthy controls, IFX=infliximab, ADA=adalimumab,

**Supplementary Table 7:**
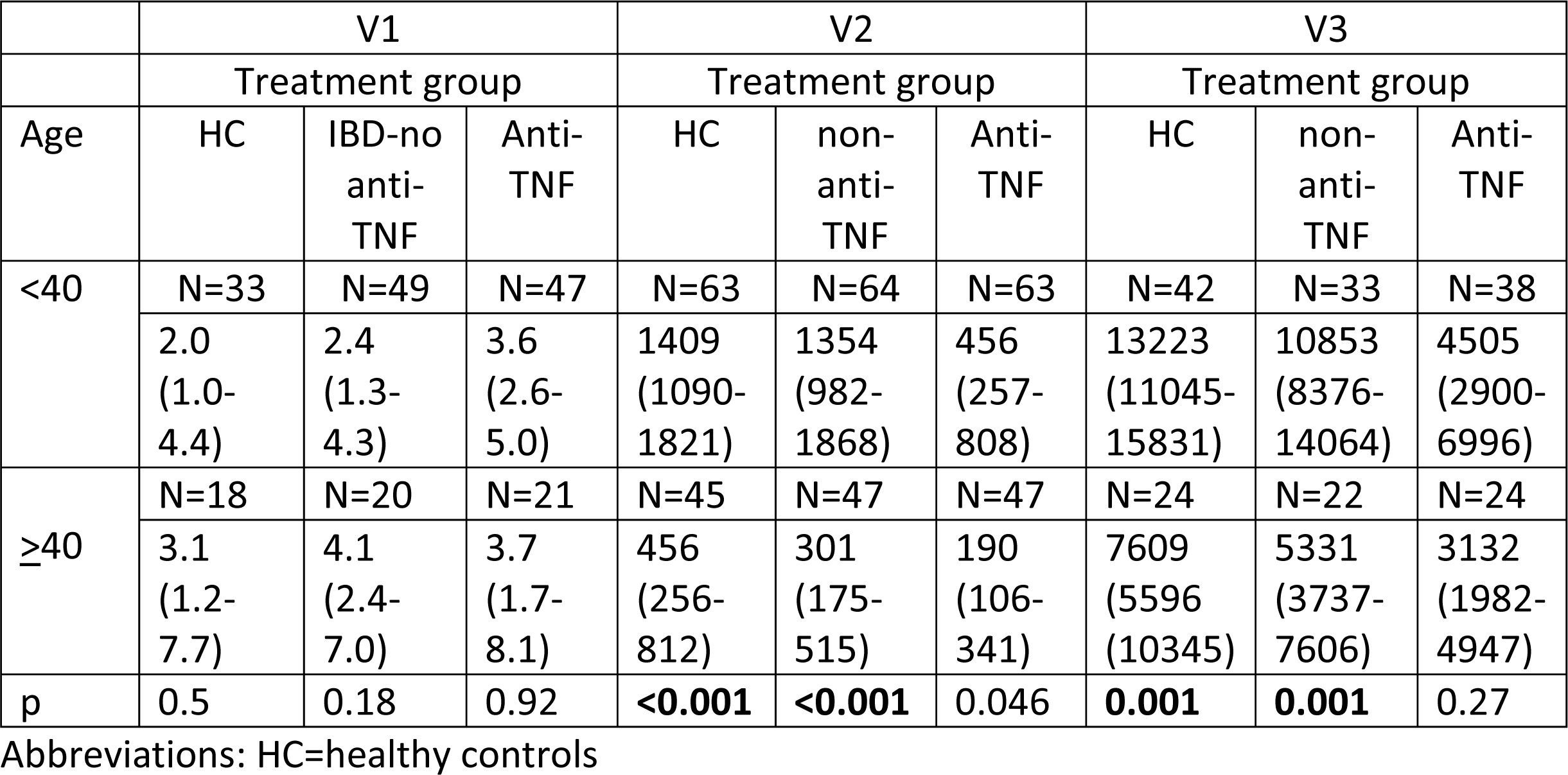
levels of IgG anti-S (GMC and 95% CI) by treatment group and age (40 years of age as cutoff) before (V1) and after first (V2) and second (V3) vaccine doses.

|  | V1 |  |  | V2 |  |  | V3 |  |  |
| --- | --- | --- | --- | --- | --- | --- | --- | --- | --- |
|  | Treatment group |  |  | Treatment group |  |  | Treatment group |  |  |
| Age | HC | IBD-no anti-TNF | Anti-TNF | HC | non-anti-TNF | Anti-TNF | HC | non-anti-TNF | Anti-TNF |
| <40 | N=33 | N=49 | N=47 | N=63 | N=64 | N=63 | N=42 | N=33 | N=38 |
|  | 2.0<br>(1.0-4.4) | 2.4<br>(1.3-4.3) | 3.6<br>(2.6-5.0) | 1409<br>(1090-1821) | 1354<br>(982-1868) | 456<br>(257-808) | 13223<br>(11045-15831) | 10853<br>(8376-14064) | 4505<br>(2900-6996) |
| ≥40 | N=18 | N=20 | N=21 | N=45 | N=47 | N=47 | N=24 | N=22 | N=24 |
|  | 3.1<br>(1.2-7.7) | 4.1<br>(2.4-7.0) | 3.7<br>(1.7-8.1) | 456<br>(256-812) | 301<br>(175-515) | 190<br>(106-341) | 7609<br>(5596-10345) | 5331<br>(3737-7606) | 3132<br>(1982-4947) |
| p | 0.5 | 0.18 | 0.92 | <0.001 | <0.001 | 0.046 | 0.001 | 0.001 | 0.27 |

Abbreviations: HC=healthy controls

**Supplementary Table 8:**
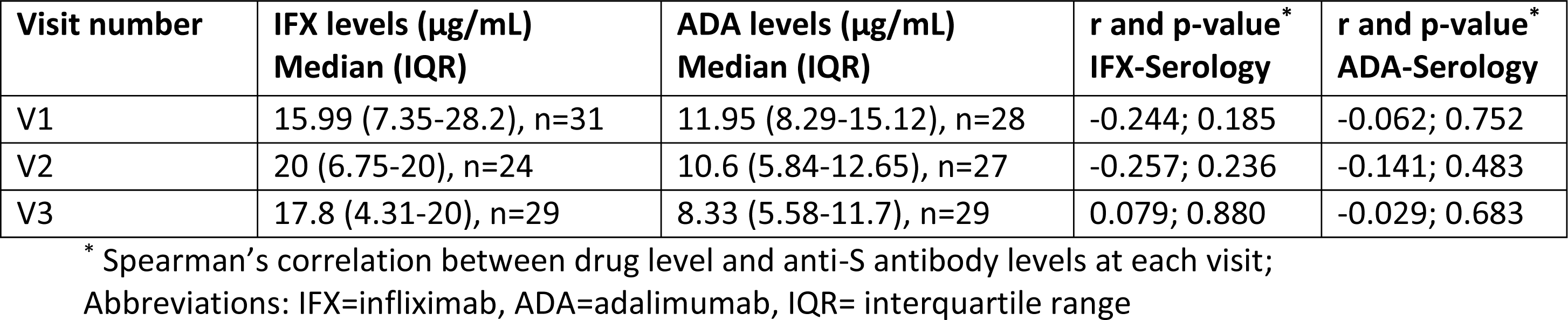
Anti-TNFα levels in each visit, and correlation with anti-S IgG levels.

| Visit number | IFX levels (µg/mL)<br>Median (IQR) | ADA levels (µg/mL)<br>Median (IQR) | r and p-value*<br>IFX-Serology | r and p-value*<br>ADA-Serology |
| --- | --- | --- | --- | --- |
| V1 | 15.99 (7.35-28.2), n=31 | 11.95 (8.29-15.12), n=28 | -0.244; 0.185 | -0.062; 0.752 |
| V2 | 20 (6.75-20), n=24 | 10.6 (5.84-12.65), n=27 | -0.257; 0.236 | -0.141; 0.483 |
| V3 | 17.8 (4.31-20), n=29 | 8.33 (5.58-11.7), n=29 | 0.079; 0.880 | -0.029; 0.683 |

\* Spearman's correlation between drug level and anti-S antibody levels at each visit; Abbreviations: IFX=infliximab, ADA=adalimumab, IQR= interquartile range

**Supplementary Table 9:**
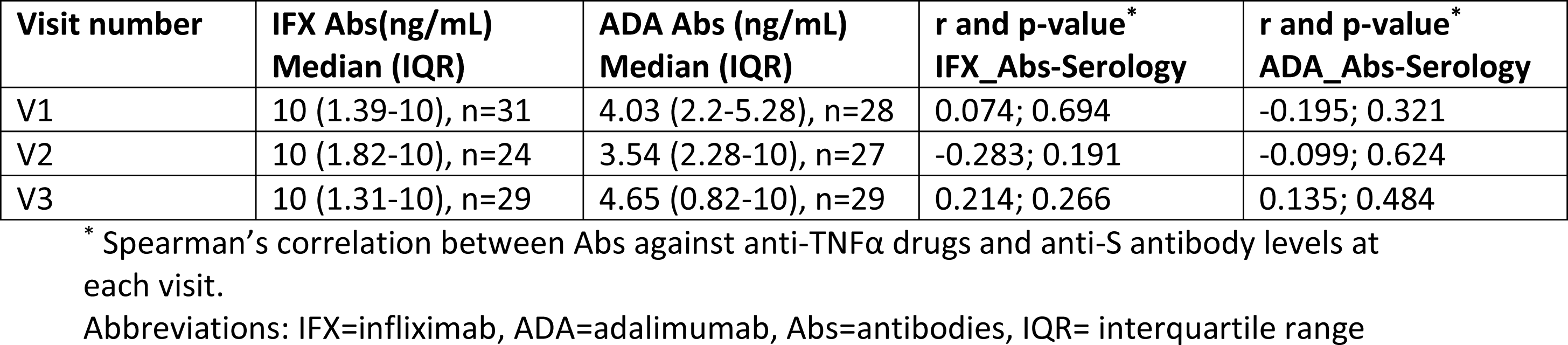
Anti-TNFα antibodies in each visit, and correlation with anti-S IgG levels.

| Visit number | IFX Abs(ng/mL)<br>Median (IQR) | ADA Abs (ng/mL)<br>Median (IQR) | r and p-value*<br>IFX_Abs-Serology | r and p-value*<br>ADA_Abs-Serology |
| --- | --- | --- | --- | --- |
| V1 | 10 (1.39-10), n=31 | 4.03 (2.2-5.28), n=28 | 0.074; 0.694 | -0.195; 0.321 |
| V2 | 10 (1.82-10), n=24 | 3.54 (2.28-10), n=27 | -0.283; 0.191 | -0.099; 0.624 |
| V3 | 10 (1.31-10), n=29 | 4.65 (0.82-10), n=29 | 0.214; 0.266 | 0.135; 0.484 |

\* Spearman's correlation between Abs against anti-TNFα drugs and anti-S antibody levels at each visit. Abbreviations: IFX=infliximab, ADA=adalimumab, Abs=antibodies, IQR= interquartile range

**Supplementary Table 10:**
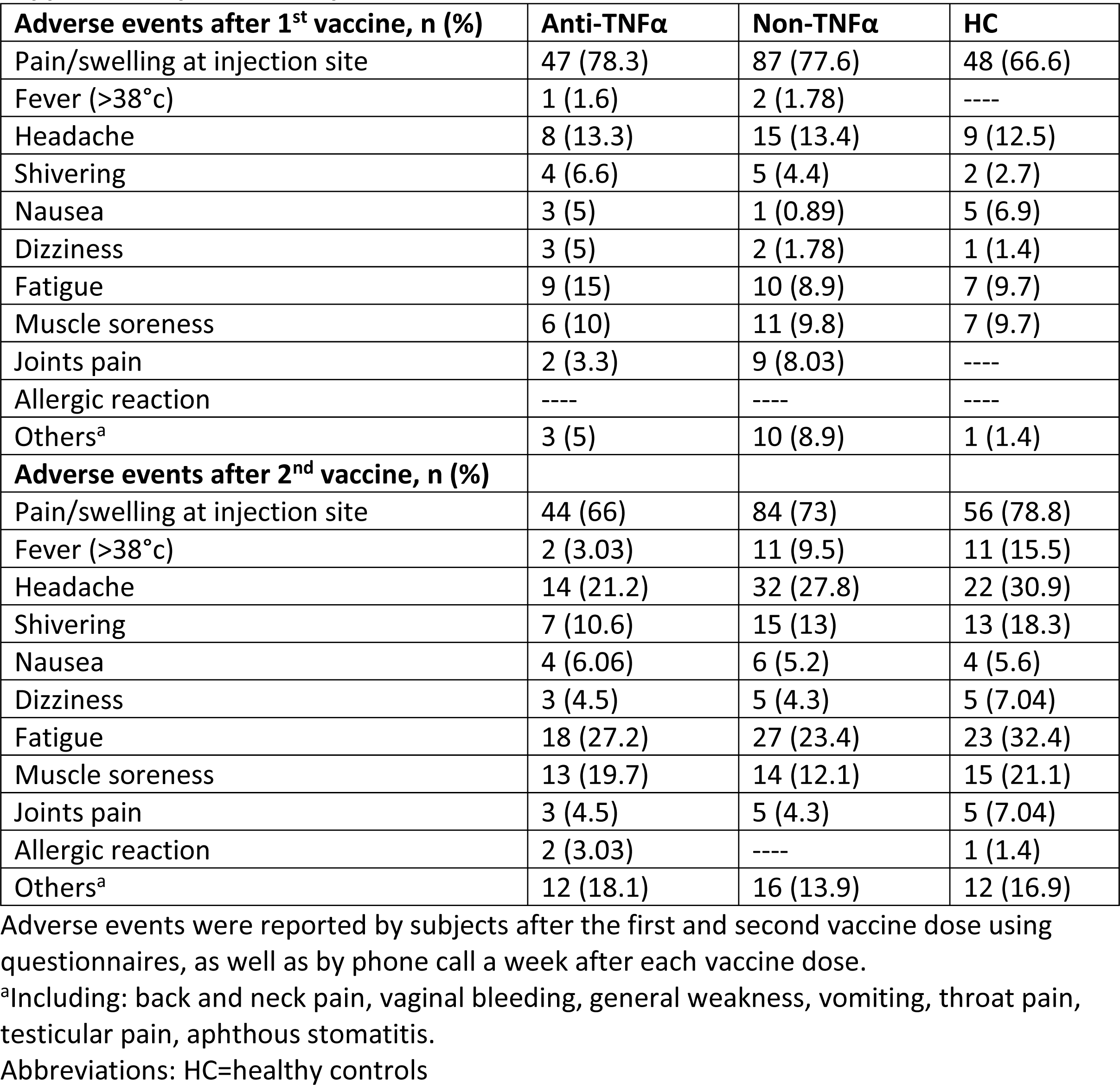
Specific adverse events.

| Adverse events after 1<sup>st</sup> vaccine, n (%) | Anti-TNFα | Non-TNFα | HC |
| --- | --- | --- | --- |
| Pain/swelling at injection site | 47 (78.3) | 87 (77.6) | 48 (66.6) |
| Fever (>38°C) | 1 (1.6) | 2 (1.78) | ---- |
| Headache | 8 (13.3) | 15 (13.4) | 9 (12.5) |
| Shivering | 4 (6.6) | 5 (4.4) | 2 (2.7) |
| Nausea | 3 (5) | 1 (0.89) | 5 (6.9) |
| Dizziness | 3 (5) | 2 (1.78) | 1 (1.4) |
| Fatigue | 9 (15) | 10 (8.9) | 7 (9.7) |
| Muscle soreness | 6 (10) | 11 (9.8) | 7 (9.7) |
| Joints pain | 2 (3.3) | 9 (8.03) | ---- |
| Allergic reaction | ---- | ---- | ---- |
| Others<sup>a</sup> | 3 (5) | 10 (8.9) | 1 (1.4) |
| Adverse events after 2<sup>nd</sup> vaccine, n (%) |  |  |  |
| Pain/swelling at injection site | 44 (66) | 84 (73) | 56 (78.8) |
| Fever (>38°C) | 2 (3.03) | 11 (9.5) | 11 (15.5) |
| Headache | 14 (21.2) | 32 (27.8) | 22 (30.9) |
| Shivering | 7 (10.6) | 15 (13) | 13 (18.3) |
| Nausea | 4 (6.06) | 6 (5.2) | 4 (5.6) |
| Dizziness | 3 (4.5) | 5 (4.3) | 5 (7.04) |
| Fatigue | 18 (27.2) | 27 (23.4) | 23 (32.4) |
| Muscle soreness | 13 (19.7) | 14 (12.1) | 15 (21.1) |
| Joints pain | 3 (4.5) | 5 (4.3) | 5 (7.04) |
| Allergic reaction | 2 (3.03) | ---- | 1 (1.4) |
| Others<sup>a</sup> | 12 (18.1) | 16 (13.9) | 12 (16.9) |

Adverse events were reported by subjects after the first and second vaccine dose using questionnaires, as well as by phone call a week after each vaccine dose.
<sup>a</sup>Including: back and neck pain, vaginal bleeding, general weakness, vomiting, throat pain, testicular pain, aphthous stomatitis.
Abbreviations: HC=healthy controls

**Supplementary Table 11:** Disease activity before and after each vaccine dose.

| V1 |  |  |  | V2 |  |  |  | V3 |  |  |  |
| --- | --- | --- | --- | --- | --- | --- | --- | --- | --- | --- | --- |
| Anti-TNFα | | Non-anti-TNFα | | Anti-TNFα | | Non-anti-TNFα | | Anti-TNFα | | Non-anti-TNFα | |
| CD<br>N=56 | UC<br>N=9 | CD<br>N=60 | UC<br>N=44 | CD<br>N=51 | UC<br>N=6 | CD<br>N=57 | UC<br>N=46 | CD<br>N=50 | UC<br>N=9 | CD<br>N=52 | UC<br>N=43 |
| 3.6±3.4 | 4.6±3.2 | 4.2±3.9 | 3.1±3.1 | 3.6±3.9 | 4.5±3.0 | 3.9±3.1 | 3.2±3.1 | 3.5±4.0 | 3.1±2.8 | 4.2±3.4 | 2.8±2.8 |

Activity was measured by validated questionnaires (HBI for patients with CD, SCCAI for patients with UC). No difference in disease activity between groups or within groups after vaccination was noticed. Data are presented as mean±SD.
Abbreviations: CD=Crohn's disease, UC=ulcerative colitis

## Supplementary Figures

**Supplementary Figure 1:**
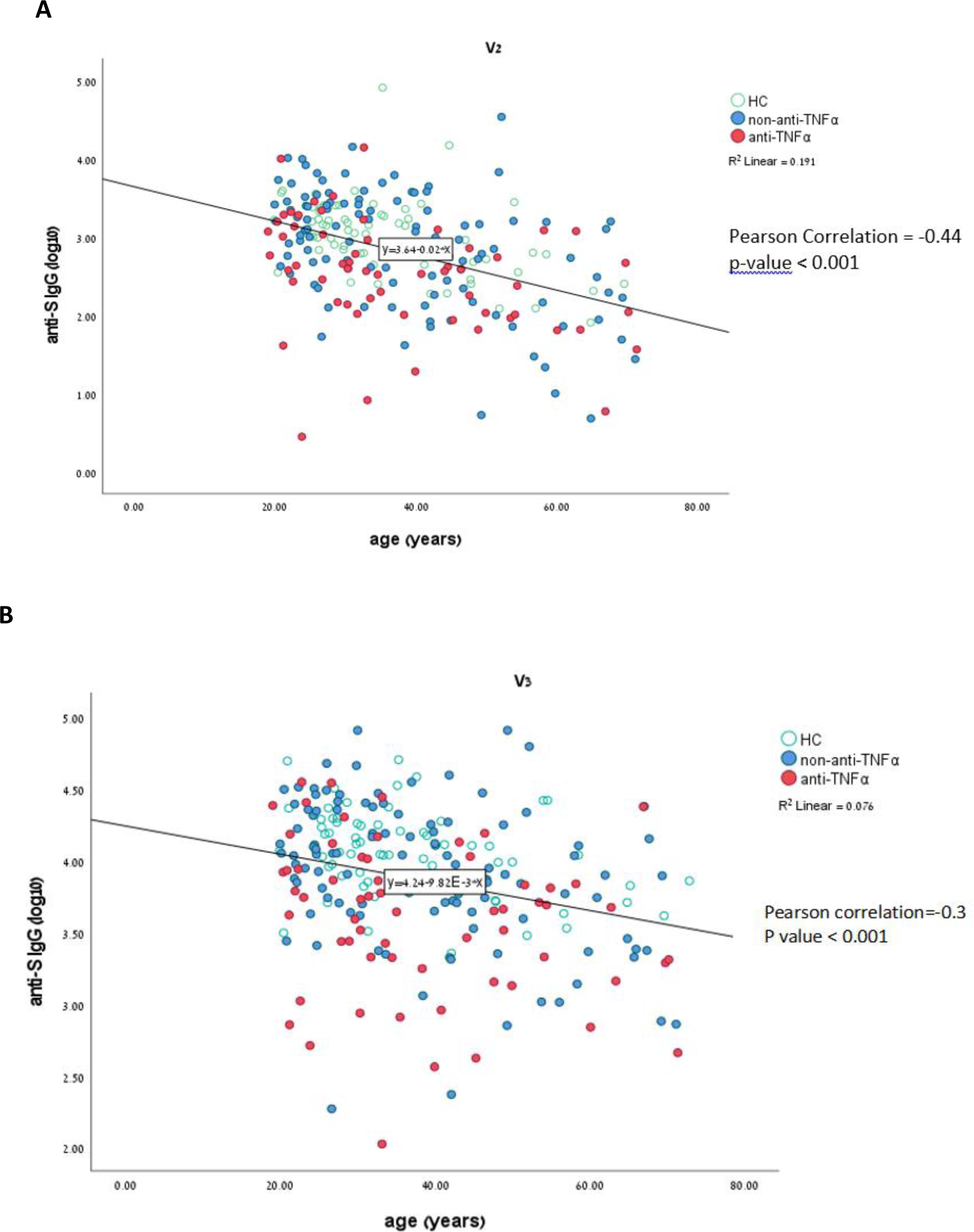
Correlation between age and serologic response. Correlation between older age and lower levels of IgG anti-S antibodies in the three study groups after first (A) and second (B) vaccine doses. Anti-TNFα in red, non-anti-TNFα in blue, healthy controls (HC) in green circles. P<0.001 in both V2 and V3 by Pearson’s correlation

**Supplementary Figure 2:**
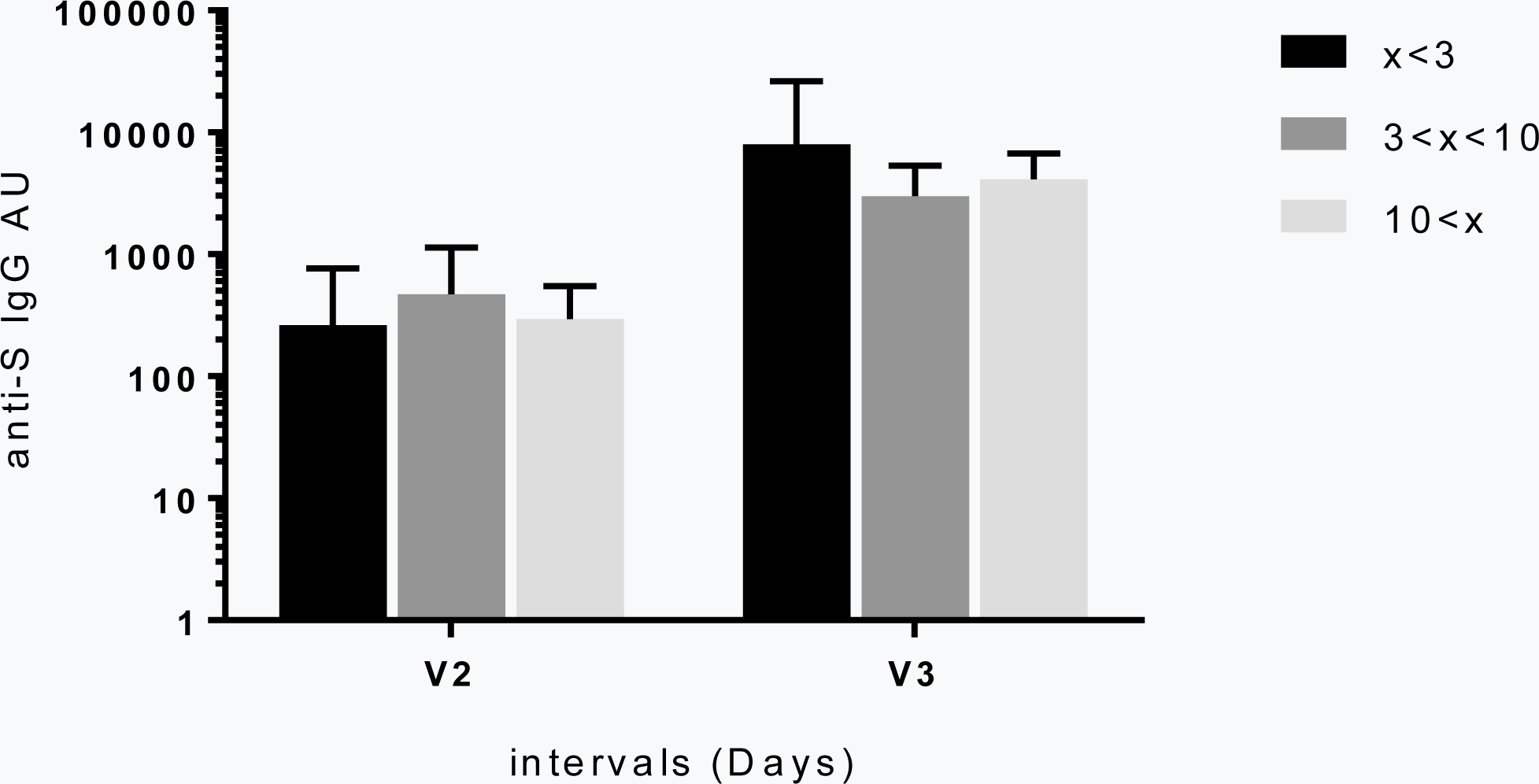
Association between serologic response and the time interval between anti-TNFα drug administration and vaccination. The bars describe the intervals in days grouped into < 3 days (black), < 10 days (dark grey), and > 10 days (light grey). Y axis: anti-S IgG antibodies after first (V2) and second (V3) vaccine doses. Error bars denote SD.

**Supplementary Figure 3:**
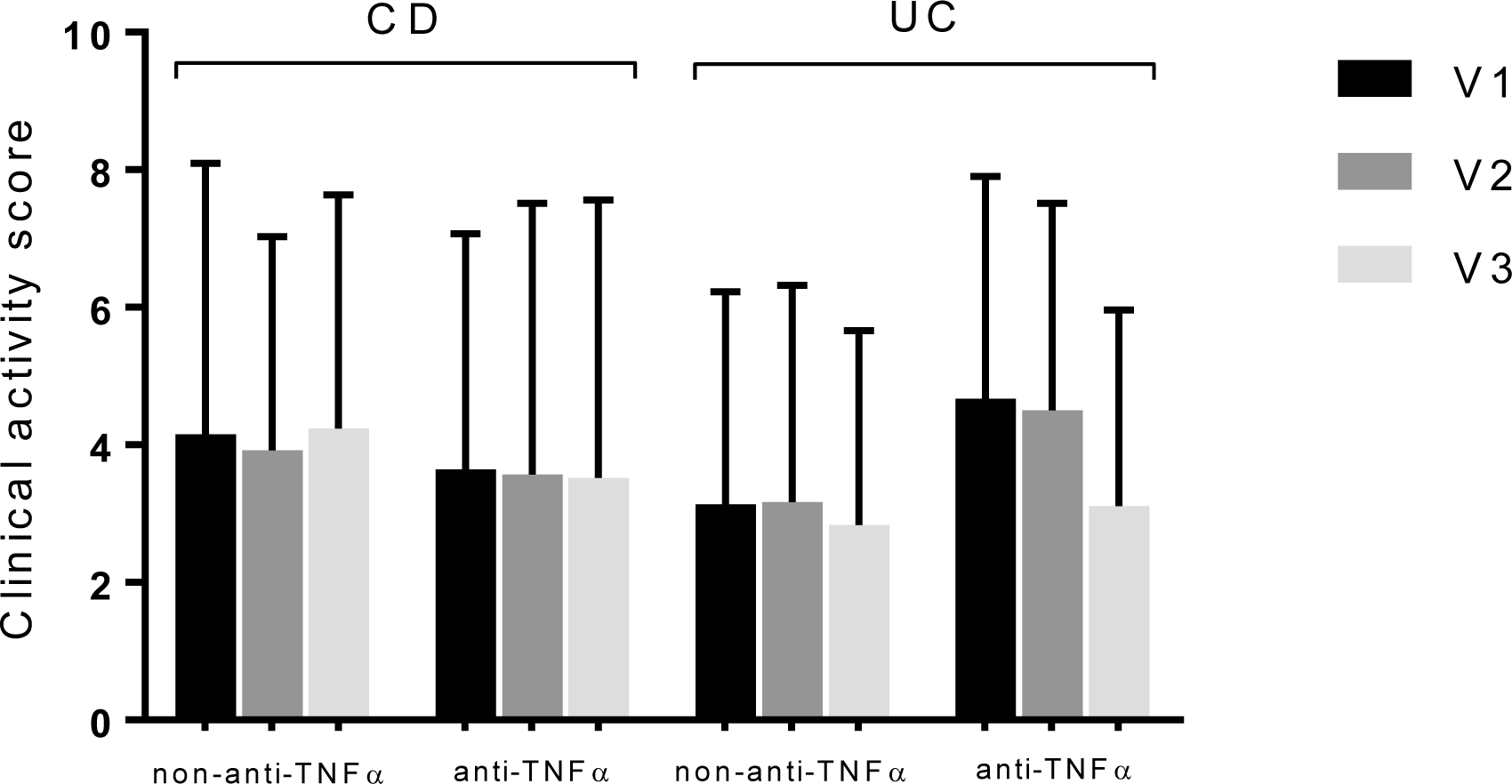
Disease activity during follow up. Activity was measured by validated questionnaires. Bars represent the average score of either HBI for CD or SCCAI for UC, stratified according to treatment (with and without anti-TNFα), before the first vaccine dose (V1, black), and before and after the second vaccine dose (V2, dark grey; V3, light grey, respectively). Error bars denote SD. Abbreviations: HBI= Harvey-Bradshaw Index; SCCAI= Simple Clinical Colitis Activity Index, UC=Ulcerative colitis, CD=Crohn’s disease

